# The Effects of Sodium Fructose Diphosphate on Blood Coagulation Reaction Time and Plasma Coagulation Factor Activity Tests in vitro

**DOI:** 10.1101/2025.02.26.25322954

**Authors:** Yalong Zhang, Xingguo Zhong, Lin Zhou, Ning He, Yuan Fang, Tongqing Chen

## Abstract

**Objective:** To investigate the effects of sodium fructose diphosphate (FDP) on blood coagulation parameters, including reaction time and plasma coagulation factor activity, in both *in vitro* and *in vivo* models.

**Methods:** Three thromboelastography systems (Maiketian, Lepu, Dingrun) were used to assess coagulation parameters (reaction time [R], clotting time [K], α-angle, maximal amplitude [MA]) in blood samples spiked with varying FDP concentrations. An automatic coagulation analyzer quantified the activities of coagulation factors II, V, VII, VIII, IX, X, XI, and XII in FDP-treated plasma.Differences between FDP-treated and control groups were statistically compared. Linear regression analyzed correlations between FDP concentrations and coagulation parameters. For *in vivo* studies, New Zealand white rabbits received intravenous FDP (0.5, 1, 2, or 4 g/kg), and thromboelastography parameters were monitored at multiple time points post-administration (0.5-2 hours).

**Results:** Thromboelastography demonstrated a strong positive correlation between FDP concentration and R values across all systems (*P* < 0.001; *r* = 0.988 [Maiketian], 0.999 [Lepu], 0.996 [Dingrun]). No significant associations were observed between FDP and K, α-angle, or MA (*P* > 0.05). *In vitro* experiments revealed significant negative correlations between FDP concentration and activities of factors V (*r* = -0.995), VII (*r* = -0.990), IX (*r* = -0.989), XI (*r* = -0.997), and XII (*r* = -0.995) (*P* < 0.001), while factors II, VIII, and X remained unaffected (*P* > 0.05). In vivo administration demonstrated dose-dependent prolongation of R-time, reaching statistical significance (p < 0.05) at:0.5 g/kg: 0.5 hr post-dose,1 g/kg: 0.5-1.5 hr,2 g/kg: 0.5-1.5 hr,4 g/kg: 0.5-2 hr.

**Conclusion:** FDP significantly impacts coagulation testing outcomes both in vitro and in vivo, potentially through modulation of intrinsic pathway factors (V, VII, IX, XI, XII) and direct interference with clot initiation. These findings suggest clinically relevant anticoagulant properties that warrant further investigation.

## 1. Introduction

Sodium fructose diphosphate (fructose-1, 6-diphosphate, FDP) is an intermediate product of intracellular glycolytic metabolism with regulatory function of key enzyme activities of glucose metabolism^1^. Exogenous FDP enters the cell through the cell membrane to activate phosphofructokinase and pyruvate kinase to increase the concentration of adenosine triphosphate and phosphocreatine in the cell, and promote the inward flow of potassium and calcium ions, which is beneficial to the energy metabolism of the cell and glucose utilization under the state of ischemia and hypoxia^2^, so as to make the ischemic tissue cells to reduce the damage, and to further improve the activity and function of the cells^3,4^. The drug began to be used in clinical practice in the 1980s, and subsequently, it has been widely used in the treatment of a variety of diseases, such as cardiovascular disease, acute adult respiratory distress syndrome, parenteral nutrition, digestive gastrointestinal disease postoperative, anemia, chronic obstructive pulmonary disease, renal insufficiency and so on^3,4^. FDP has a wide range of pharmacological effects, from the molecular level to participate in the regulation of a variety of intracellular metabolic processes, thereby improving cellular energy metabolism, increasing energy utilization, inhibiting the generation of free radicals, maintaining cell membrane stability, accelerating tissue repair, and maintaining organ function^4,5^. A large number of studies have confirmed that FDP can be used individually or adjunctively in the treatment of various causes of tissue ischemia, hypoxia and organ damage^5^, exogenous FDP has almost no toxic side effects in the process of drug administration, and there is almost no contraindication, can be administered orally or intravenously, and has been widely used in the clinical environment, which has shown good social and economic benefits.

Although a large number of studies have shown that the use of FDP in the treatment of many diseases is efficacious and has few toxic side effects, the side effects of this drug are still not well understood and have only been sporadically reported, and so far, even less is known about whether this drug has any effect on the coagulation process. Thromboelastography (TEG) is a sensitive test used to reflect the coagulation process in whole blood, allowing a comprehensive assessment of platelet function, plasma factor activity, fibrin polymerization and fibrinolysis^6,7^. In this article, we further explored and clarified whether FDP has an effect on the blood coagulation process and on the activities of its coagulation factors (II, V, VII, VIII, IX, X, Ⅺ, Ⅻ) through in vitro experiments using TEG and coagulation factor activity monitoring.

## 2. Methods

### 2.1 Instruments and reagents

TEG analyzer and corresponding reagents from three different manufacturers ( Maketian, Lepu and Dinrun ) were used to analyze the blood sample coagulation reaction time (R), clotting formation time (K), coagulation Angle (α-Angle) and maximum amplitude (MA). The Maiketian TEG instrument reagent testing system is based on the Maiketian Haema TX Thromboelastography Test System and its corresponding reagents(Lot 20231101, Shenzhen Maiketian Biomedical Technology Co. Ltd.). LEPU CFMS LEPU-8880 Thromboelastography Analyzer and its reagents (Thrombelastograph General Cup Test Kit viscosity measuring, Lot 23SH0102.) were provided by Lepu Medical Technology Co. Ltd. Dingrun DRNX-Ⅲ Thrombelastograph Analyzer and its supporting reagents (Activated Coagulation Reagent, Lot 20230504) were manufactured by Chongqing Dingrun Medical Equipment Co. Ltd. The Sysmex Coagulation Testing System is based on the Sysmex CS5100 Coagulation Analyzer and its corresponding reagents, including Dade Actin Activated Cephaloplastin reagent for aPTT testing (Lot 562729A), Thromborel S for PT (Lot 568182), Coagulation Factor II Deficient Plasma (Lot 503659), Coagulation Factor V Deficient Plasma (Lot: 575712), Coagulation Factor Ⅶ Deficient Plasma (Lot 500776), Coagulation Factor VIII Deficient Plasma (Lot: 560857A), Coagulation Factor IX Deficient Plasma (Lot: 504172B), Coagulation Factor X Deficient Plasma (Lot 504029), Coagulation Factor Ⅺ Deficient Plasma (Lot 503358B), Coagulation Factor Ⅻ Deficient Plasma (Lot 503427), Standard Human Plasma (Lot 563120), CONTROL N (Lot 507936), CONTROL P (Lot 556743), and the reagents were provided by Siemens Medical Diagnostic Products GmbH, Germany. Sodium fructose diphosphate was purchased from Anhui Weilman Pharmaceutical Co. Ltd, China (Lot 20231001).

### 2.2 Sample testing methods and procedures

The following experimental procedures and protocols were approved by the Ethics Review Committee of Anhui No.2 Provincial People’s Hospital, China [(R) 2024-037]. Prior to this test, all assay systems, except for normal calibration, were performed using commercially available control samples specified by the manufacturer of each assay system supporting traceability, including normal and abnormal quality control samples. Each sample was then tested on the machine with the assay system under normal conditions.

### 2.3 Blood sample collection and preparation

About 14 ml blood sample (no history of blood disorders such as platelet and coagulation disorders, and no medication that affects coagulation, such as aspirin, in the past 2 weeks) was randomly collected from 11 volunteers veins for normal physical health examination, sample anticoagulated by trisodium citrate at 1.09 mmol/L (blood-to-citric-acid anticoagulation ratio of 9:1).The recruitment period for the study was from April 15 to May 15, 2024.In this study, all participants signed a written informed consent, and we also orally informed them of the specific research content. The study did not involve minors. Informed consent terms and contents are subject to approval by the hospital Ethics Committee.In this study, samples were randomly changed by double-blind method after collection. During or after data collection, the authors were not able to obtain information that would identify individual participants.

### 2.4 Preparation and detection of thromboelastographic samples

Collect 7 mL of anticoagulant blood samples from each of the aforementioned subjects, along with 7 polyethylene graduated centrifuge tubes. Pipette 1 mL of the blood sample into each tube, and precisely weigh a certain quantity of FDP using an analytical balance. Subsequently, add the weighed FDP into the tubes, ensuring that the final FDP concentrations in the blood - FDP mixtures are 0 (control tube), 1, 2, 3, 4, 5, and 6 mg/mL respectively. Repeat this procedure for the other 10 mixed blood samples. In total, prepare 11 blood samples obtained from the veins of different volunteers. Test the coagulation reaction time (R), clotting time (K), α-angle (α-Angle), and maximal amplitude (MA) using the Maiketian, Lepu, and Dingrun thromboelastography systems respectively.

### 2.5 Preparation and testing of coagulation factor plasma samples

Take 7 mL of anticoagulant blood samples from each of the subjects mentioned above and 7 polyethylene graduated centrifuge tubes. Add 1 mL of blood sample to each tube. Centrifuge the tubes horizontally at 3000 r/min for 10 min with a radius of R = 15 cm to obtain approximately 0.5 mL of plasma samples containing coagulation factors. Then, add a specific amount of FDP to each plasma tube, so that the final FDP concentrations in the plasma - FDP mixtures are 0 (control tube), 1, 2, 3, 4, 5, and 6 mg/mL respectively. Conduct the same operation for the other 10 mixed plasma samples. After gentle and thorough mixing, measure the activities of coagulation factors II, V, VII, VIII, IX, X, Ⅺ, and Ⅻ in each mixed plasma sample using a Sysmex CS5100 fully automated blood coagulation detection analyzer.

### 2.6 Animal experiments

A total of 30 healthy adult male New Zealand Large White rabbits, weighing between 2.3 and 3.0 kg, were procured from the Laboratory Animal Center of Anhui Medical University under the license No. SYXK(Wan)2023 - 015. All experimental procedures strictly adhered to "Guidelines for Ethical Review of Animal Welfare (GB/T 35892 - 2018, China)" (version 2.0) and were approved by the Animal Experiment Ethics Committee of Anhui University of Chinese Medicine (AHUCM - rabbit - 20240051).

The rabbits were housed individually in cages with a 12 h/12 h light/dark cycle. They were provided with free access to food and water at a room temperature of around 25°C and a relative humidity of approximately 50% for one week to allow them to acclimate to the environment. Before each experiment, the rabbits were fasted for 4 - 6 hours while having free access to water. Randomly divide the thirty New Zealand White rabbits into 5 groups: a control group receiving 0.0 g/kg FDP (in the form of saline injection) and four experimental groups receiving 0.5 g/kg, 1 g/kg, 2 g/kg, and 4 g/kg FDP respectively, with 6 rabbits in each group. If a normal dose of 10 g FDP is injected into a Chinese person weighing 70 kg, the equivalent intravenous dose for a domestic rabbit weighing 2.5 kg is approximately 0.467 g/kg (calculated as 10/70×3.27, a rough conversion based on body surface area). Inject the corresponding doses of FDP into the experimental groups intravenously through the rabbit ear margins within one minute, while injecting an equal volume of saline into the control group.

Collect 1 mL of blood samples from the central artery of the ear before FDP administration and at 0.5 h, 1 h, 1.5 h, 2 h, and 3 h after administration. Place the samples in polyethylene vacuum blood collection tubes containing 3.8% sodium citrate at a ratio of 1:9 (sodium citrate:blood). After thorough mixing and anticoagulation, complete the detection of R, K, α - angle, and MA using the Maiketian thromboelastography system within 4 hours.

#### Anesthesia and Analgesia

##### Pre - Anesthesia Preparation

Fasting: Withhold food for 4 - 6 hours while allowing free access to water to prevent aspiration.

Acclimatization: House rabbits in a quiet, temperature - controlled environment (20 - 24°C) for at least 7 days before the experiment.

##### Anesthesia Protocol

Induction: Administer ketamine (35mg/kg) and xylazine (5mg/kg) intramuscularly (IM) for sedation.

Maintenance: Use isoflurane (1- 3%) via a facemask to maintain anesthesia. Continuously monitor vital signs, including respiratory rate, heart rate, and SpO₂.

Local Anesthesia: Apply 2% lidocaine gel at the venipuncture site to minimize pain from needle insertion. Provide soft bedding and environmental enrichment (such as hiding spaces) to reduce stress.

##### Refinement Strategies to Alleviate Suffering

Blood Sampling Technique: Use a catheter placed in the marginal ear vein to avoid repeated needle insertions.

Monitoring: Assess rabbits hourly for signs of pain (e.g., vocalization, guarding behavior).

Fluid Support: Administer warm saline (10 mL/kg subcutaneously) after the procedure to prevent dehydration.

##### Euthanasia Method

Agent: Administer an overdose of sodium pentobarbital (150 mg/kg IV).

Confirmation: Verify death by the absence of a heartbeat, respiratory arrest, and fixed dilated pupils.

Disposal: Handle carcasses according to institutional biosafety protocols.

### 2.7 Statistical analysis

Prior to testing, the manufacturer validated the performance of each assay system, which successfully passed the performance validation. According to the manufacturer’s protocol, test each sample in three independent replicates and complete the testing within two hours. Take the test results of samples with an FDP concentration of 0 mg/mL as control tubes, and calculate the percentage change between the test results of each sample and the control tube results. This enables us to infer, based on the degree of change, whether the drugs have an impact on the coagulation test results and to further assess the presence of concentration - dependent interferences in the coagulation tests.

Perform all statistical analyses using Microsoft Excel 2003 (Microsoft Corporation, Redmond, WA, USA). Conduct linear regression analyses using Statistical Products and Services Solutions 20.0 (SPSS20.0, IBM, Armonk, USA) statistical software. Use the Spearman Correlation coefficients to assess the correlation coefficients between the FDP concentration and each of the R, K, α - Angle, MA values, or the activities of coagulation factors II, V, VII, VIII, IX, X, Ⅺ, and Ⅻ.

To obtain linear regression equations, conduct statistical hypothesis tests on the regression coefficients to determine the presence or absence of a linear relationship. Use P < 0.05 as the criterion to determine statistical significance. Consider the plasma or blood samples with 0 mg/mL FDP as the corresponding control groups, and use P< 0.05 to determine if there is a significant difference.

## 3. Results

### 3.1 Correlation Between FDP Concentration and Thromboelastographic Parameters

Correlation analyses revealed a strong positive association (p<0.001) between fibrin degradation product (FDP) concentration and thromboelastography (TEG)-derived coagulation reaction time (R value) across three commercial systems: Makotian (r=0.988), Lepu (r=0.999), and Dinrun (r = 0.996). Notably, no significant correlations were observed between FDP levels and other TEG parameters including clot formation time (K), α -angle, or maximum amplitude (MA) (Table 1; Figures 1-3).

**Table 1.** The coagulation time (R, min), coagulation time (K, min), maximum amplitude (MA,mm), and alpha angle(deg) of the pooled blood containing with different concentrations FDP in vitro detected by the Maiketian, Lepu, and Dingrun thromboelastography analysis systems, respectively (x̄±s, n=11) :\**p*<0.000, There is a significant difference in statistical results compared to the control group (0mg/ml FDP).Abbreviations:FDP,Fructose diphosphate.

| FDP<br>(mg/mL<br>) | Maiketian |  |  |  | Lepu |  |  |  | Dingrun |  |  |  |
| --- | --- | --- | --- | --- | --- | --- | --- | --- | --- | --- | --- | --- |
|  | R(min) | K(min) | Angle(deg) | MA(mm) | R(min) | K(min) | Angle(deg) | MA(mm) | R(min) | K(min) | Angle(deg) | MA(mm) |
| 0 | 5.66±<br>0.49 | 2.52±0.27 | 67.35±2.76 | 63.51±2.28 | 6.90±0.46 | 2.20±0.26 | 68.14±2.53 | 63.8±2.62 | 6.65±0.36 | 2.05±0.27 | 69.37±2.51 | 64.14±2.43 |
| 1 | 6.53±<br>0.50* | 2.41±0.31 | 68.75±2.73 | 64.83±3.07 | 7.30±0.31* | 2.21±0.26 | 68.47±2.00 | 64.33±2.85 | 7.29±0.49* | 2.18±0.28 | 69.81±2.31 | 65.08±2.67 |
| 2 | 6.9±<br>0.51* | 2.37±0.28 | 67.89±2.48 | 64.84±2.88 | 7.88±0.38* | 2.20±0.30 | 68.25±2.45 | 64.03±2.88 | 7.84±0.59* | 2.16±0.27 | 70.05±2.44 | 64.23±2.52 |
| 3 | 7.16±<br>0.55* | 2.41±0.26 | 67.72±2.42 | 64.85±2.79 | 8.41±0.45* | 2.27±0.27 | 67.93±2.27 | 64.20±2.53 | 8.31±0.61* | 2.22±0.30 | 70.18±2.58 | 64.38±2.43 |
| 4 | 7.55±<br>0.43* | 2.37±0.20 | 68.08±1.78 | 65.00±2.73 | 8.79±0.47* | 2.24±0.32 | 68.18±2.22 | 64.26±2.85 | 8.75±0.63* | 2.11±0.25 | 69.84±2.14 | 64.81±2.48 |
| 5 | 8.24±<br>0.49* | 2.44±0.29 | 67.51±1.88 | 64.91±2.82 | 9.32±0.46* | 2.22±0.33 | 68.45±2.07 | 64.44±2.74 | 9.48±0.63* | 2.16±0.25 | 69.87±2.44 | 64.64±2.69 |
| 6 | 8.51±<br>0.47* | 2.53±0.30 | 66.92±2.22 | 63.70±3.10 | 9.86±0.45* | 2.25±0.32 | 68.22±2.27 | 64.16±2.52 | 10.22±0.63* | 2.18±0.25 | 69.57±2.13 | 64.15±2.44 |
| *r* | 0.988 | 0.106 | -0.474 | 0.109 | 0.999 | 0.714 | 0.054 | 0.563 | 0.996 | 0.417 | 0.144 | -0.057 |
| Regres<br>sion<br>equati<br>on | $Y_{result}=0.4507X_{FDP}+5.8693$ | $Y_{result}=0.0032X_{FDP}+2.4261$ | $Y_{result}=-0.1279X_{FDP}+68.129$ | $Y_{result}=0.0318X_{FDP}+64.425$ | $Y_{result}=0.4939X_{FDP}+6.8696$ | $Y_{result}=0.0075X_{FDP}+2.2046$ | $Y_{result}=0.0046X_{FDP}+68.22$ | $Y_{result}=0.0546X_{FDP}+64.01$ | $Y_{result}=0.5714X_{FDP}+6.6486$ | $Y_{result}=0.0107X_{FDP}+2.1193$ | $Y_{result}=0.0182X_{FDP}+69.758$ | $Y_{result}=-0.0096X_{FDP}+64.519$ |
| *P* | 0.000 | 0.822 | 0.282 | 0.816 | 0.000 | 0.071 | 0.908 | 0.188 | 0.000 | 0.352 | 0.759 | 0.903 |

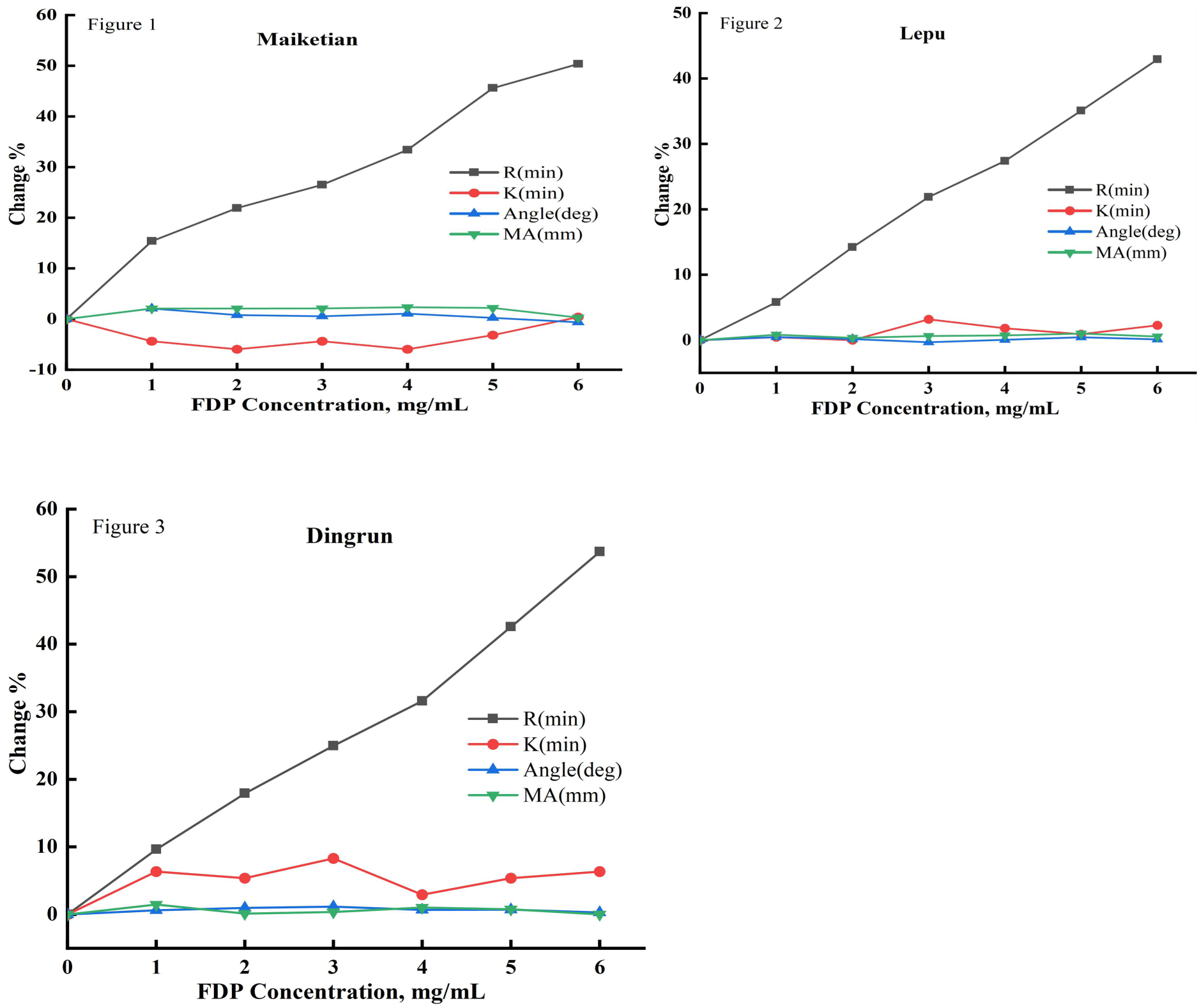
Figure 1, Figure 2 and Figure 3. Abbreviations:FDP, fructose diphosphate; Maiketian, Maiketian TX thromboelastography analyzer detection system; Lepu, Lepu CFMS LEPU-8880 thromboelastography analyzer detection system:Dingrun, Dingrun DRNX-III thromboelastography detection system. The thromboelastography coagulation time (R, min), coagulation time (K, min), maximum amplitude (MA,mm), and alpha angle(Angle) of the pooled blood after mixed with different concentrations FDP in vitro dected respectively by the Maiketian (Figure 1), Lepu (Figure 2), and Dingrun (Figure 3) thromboelastography analysis systems. The results of R were increased in a FDP concentration (0-6mg/mL) dependent way after the addition of FDP in the mixed blood in vitro. All three detection systems showed that as the concentration of FDP increased, the R results of sample showed a dependence on the increase of FDP concentration (0-6mg/mL). The Maiketian detection system showed an increase of 0∼50.35%, Lepu 0∼42.9% and Dingrun 0∼53.68%, while the result of k, MA, and Angle showed no significant changes. The R value increase with the increase of FDP concentration in blood samples, showing a linear positive correlation trend.

### 3.2 Dose-Dependent Inhibition of Coagulation Factors by FDP

In vitro studies demonstrated concentration-dependent suppression of factors V (r=-0.995), VII (r =-0.990), IX (r=-0.989), XI (r=-0.997), and XII (r=-0.995) activity with increasing FDP concentrations (0-6 mg/mL; p< 0.001). In contrast, factors II, VIII, and X exhibited no statistically significant response to FDP exposure (p> 0.05) (Table 2; Figures 4-5).

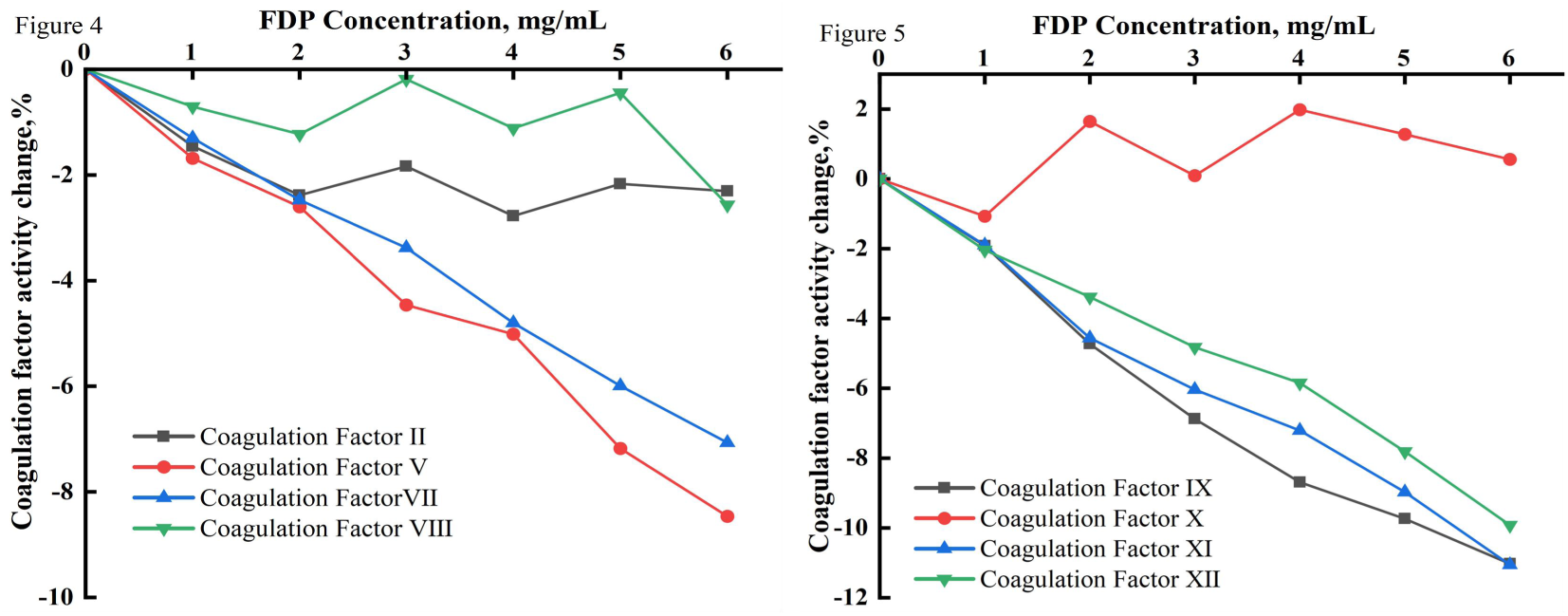
Figure 4. and Figure 5. The mean change with the activity for plasma coagulation factors (II, V, VII, VIII, IX, X, Ⅺ, Ⅻ) of mixed blood with different concentrations of FDP added was determined by Sysmex CS5100 Coagulation Analyzer System (x̄±s, n=11). With the increase of FDP concentration (0, 1, 2, 3, 4, 5 and 6mg/mL),the activities of Coagulation factor Ⅴ, Ⅶ, Ⅸ, Ⅺ and Ⅻ showed a significant downward trend(*p*<0.000), while Ⅱ,Ⅷ and Ⅹ showed no significant statistical changes. Abbreviations: FDP, Fructose diphosphate

**Table 2.**
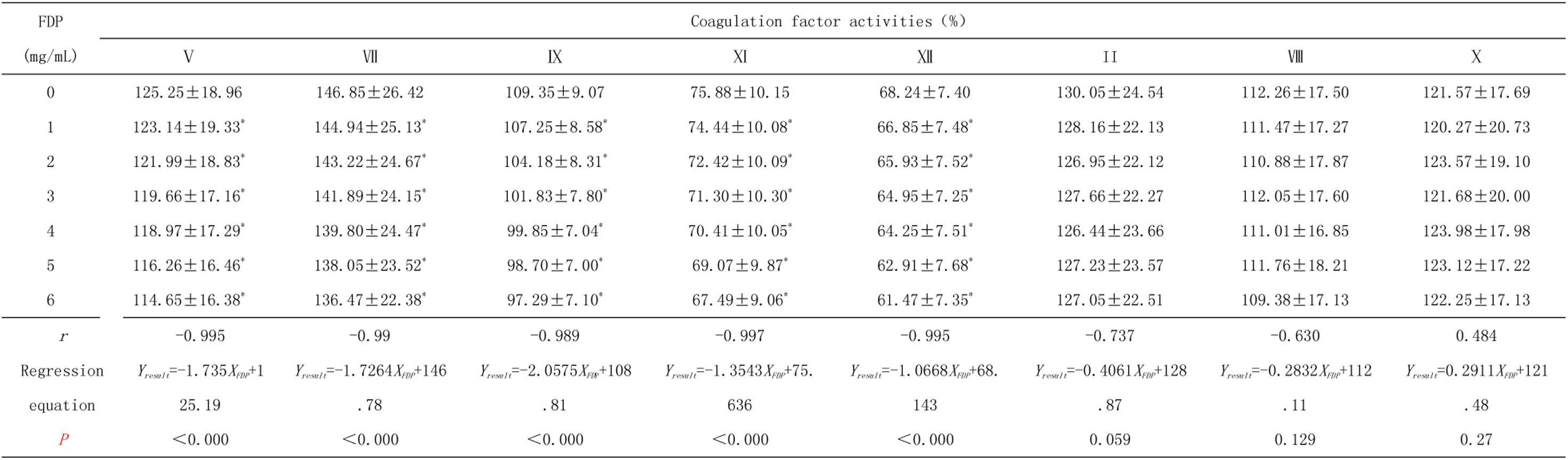
Coagulation factor (Ⅱ、Ⅴ、Ⅶ、Ⅷ、Ⅸ、Ⅹ、Ⅺ、Ⅻ) activities (%) of the pooled plasma containing with different concentrations FDP in vitro detected by Sysmex CS5100 coagulation analysis system (x̄±s, n=11) :*p<0.000, There is a significant difference in statistical results compared to the control group (0mg/ml FDP).Abbreviations:FDP,fructose diphosphate.

| FDP<br>(mg/mL) | Coagulation factor activities (%) |  |  |  |  |  |  |  |
| --- | --- | --- | --- | --- | --- | --- | --- | --- |
|  | V | VII | IX | XI | XII | II | VIII | X |
| 0 | 125.25±18.96 | 146.85±26.42 | 109.35±9.07 | 75.88±10.15 | 68.24±7.40 | 130.05±24.54 | 112.26±17.50 | 121.57±17.69 |
| 1 | 123.14±19.33* | 144.94±25.13* | 107.25±8.58* | 74.44±10.08* | 66.85±7.48* | 128.16±22.13 | 111.47±17.27 | 120.27±20.73 |
| 2 | 121.99±18.83* | 143.22±24.67* | 104.18±8.31* | 72.42±10.09* | 65.93±7.52* | 126.95±22.12 | 110.88±17.87 | 123.57±19.10 |
| 3 | 119.66±17.16* | 141.89±24.15* | 101.83±7.80* | 71.30±10.30* | 64.95±7.25* | 127.66±22.27 | 112.05±17.60 | 121.68±20.00 |
| 4 | 118.97±17.29* | 139.80±24.47* | 99.85±7.04* | 70.41±10.05* | 64.25±7.51* | 126.44±23.66 | 111.01±16.85 | 123.98±17.98 |
| 5 | 116.26±16.46* | 138.05±23.52* | 98.70±7.00* | 69.07±9.87* | 62.91±7.68* | 127.23±23.57 | 111.76±18.21 | 123.12±17.22 |
| 6 | 114.65±16.38* | 136.47±22.38* | 97.29±7.10* | 67.49±9.06* | 61.47±7.35* | 127.05±22.51 | 109.38±17.13 | 122.25±17.13 |
| *r* | -0.995 | -0.99 | -0.989 | -0.997 | -0.995 | -0.737 | -0.630 | 0.484 |
| Regression equation | $Y_{result} = -1.735X_{FDP} + 125.19$ | $Y_{result} = -1.7264X_{FDP} + 146.78$ | $Y_{result} = -2.0575X_{FDP} + 108.81$ | $Y_{result} = -1.3543X_{FDP} + 75.636$ | $Y_{result} = -1.0668X_{FDP} + 68.143$ | $Y_{result} = -0.4061X_{FDP} + 128.87$ | $Y_{result} = -0.2832X_{FDP} + 112.11$ | $Y_{result} = 0.2911X_{FDP} + 121.48$ |
| *P* | <0.000 | <0.000 | <0.000 | <0.000 | <0.000 | 0.059 | 0.129 | 0.27 |

### 3.3 Temporal Effects of Intravenous FDP on Coagulation Kinetics

Longitudinal thromboelastographic monitoring in New Zealand white rabbits revealed dose- and time-dependent prolongation of R values following FDP administration (0.5-2 g/kg):0.5 g/kg: Significant R value increase at 0.5 hr post-injection (p<0.01);1 g/kg: Sustained effects at 0.5, 1, and 1.5 hr (p<0.01);2 g/kg: Prolonged effects persisting through 2 hr (p< 0.01);No significant alterations were observed in K, α-angle, or MA parameters across dose groups or timepoints (p > 0.05) (Table 3; Figure 6).

**Table 3.** Thromboelastograms (R, K, α-angle, and MA) of different dose groups at different point time before and after FDP intravenous injection (x̄±s, n=6)

| time (h) | R (min) |  |  |  |  | F | P |
| --- | --- | --- | --- | --- | --- | --- | --- |
|  | 0g/Kg group | 0.5g/Kg group | 1g/Kg group | 2g/Kg group | 4g/Kg group |  |  |
| 0<sup>a</sup> | 3.52±0.21 | 3.67±0.23 | 3.62±0.12 | 3.50±0.14 | 3.67±0.12 | 1.38 | 0.26 |
| 0.5<sup>b</sup> | 3.47±0.23 | 4.23±0.23* | 5.45±0.21* | 6.65±0.23* | 12.43±0.32* | 1241.1 | 0.000 |
| 1<sup>b</sup> | 3.47±0.15 | 3.93±0.33 | 6.08±0.19* | 7.65±0.23* | 9.60±0.30* | 643.8 | 0.000 |
| 1.5<sup>b</sup> | 3.42±0.28 | 3.78±0.31 | 4.53±0.15* | 5.35±0.19* | 5.85±0.16* | 122.1 | 0.000 |
| 2<sup>b</sup> | 3.48±0.23 | 3.62±0.21 | 3.58±0.15 | 3.58±0.15 | 4.32±0.20* | 18.7 | 0.000 |
| 3<sup>b</sup> | 3.38±0.15 | 3.77±0.24 | 3.62±0.16 | 3.52±0.20 | 3.62±0.19 | 3.27 | 0.027 |

| time (h) | K (min) |  |  |  |  | F | P |
| --- | --- | --- | --- | --- | --- | --- | --- |
|  | 0g/Kg group | 0.5g/Kg group | 1g/Kg group | 2g/Kg group | 4g/Kg group |  |  |
| 0<sup>a</sup> | 1.32±0.12 | 1.38±0.10 | 1.42±0.17 | 1.47±0.16 | 1.48±0.15 | 1.33 | 0.286 |
| 0.5<sup>b</sup> | 1.42±0.15 | 1.43±0.12 | 1.45±0.19 | 1.46±0.20 | 1.37±0.18 | 0.27 | 0.893 |
| 1<sup>b</sup> | 1.38±0.15 | 1.33±0.14 | 1.45±0.16 | 1.57±0.14 | 1.50±0.24 | 1.75 | 0.171 |
| 1.5<sup>b</sup> | 1.38±0.16 | 1.42±0.17 | 1.48±0.19 | 1.53±0.21 | 1.48±0.21 | 0.83 | 0.518 |
| 2<sup>b</sup> | 1.33±0.16 | 1.33±0.12 | 1.43±0.14 | 1.40±0.17 | 1.55±0.22 | 1.78 | 0.165 |
| 3<sup>b</sup> | 1.32±0.12 | 1.35±0.10 | 1.35±0.19 | 1.48±0.15 | 1.47±0.19 | 1.49 | 0.234 |

| time (h) | α-angle (deg) |  |  |  |  | F | P |
| --- | --- | --- | --- | --- | --- | --- | --- |
|  | 0g/Kg group | 0.5g/Kg group | 1g/Kg group | 2g/Kg group | 4g/Kg group |  |  |
| 0<sup>a</sup> | 76.05±2.14 | 77.18±2.57 | 75.82±2.12 | 75.08±2.47 | 76.83±2.22 | 0.78 | 0.547 |
| 0.5<sup>b</sup> | 76.82±2.54 | 77.35±0.23 | 74.83±1.83 | 75.97±2.15 | 76.30±1.91 | 0.86 | 0.504 |
| 1<sup>b</sup> | 75.78±1.65 | 76.17±2.53 | 74.80±1.38 | 75.58±2.40 | 76.72±2.13 | 0.71 | 0.592 |
| 1.5<sup>b</sup> | 75.02±1.60 | 76.27±2.81 | 75.32±2.04 | 75.77±1.95 | 75.38±2.73 | 0.27 | 0.896 |
| 2<sup>b</sup> | 74.55±1.81 | 75.88±2.15 | 74.67±1.91 | 76.63±2.26 | 75.85±2.34 | 1.06 | 0.394 |
| 3<sup>b</sup> | 76.28±2.07 | 76.63±2.20 | 75.43±2.54 | 76.88±2.17 | 75.78±2.68 | 0.32 | 0.863 |

| time (h) | MA (mm) |  |  |  |  | F | P |
| --- | --- | --- | --- | --- | --- | --- | --- |
|  | 0g/Kg group | 0.5g/Kg group | 1g/Kg group | 2g/Kg group | 4g/Kg group |  |  |
| 0<sup>a</sup> | 73.13±1.92 | 72.22±2.27 | 75.52±2.61 | 71.47±2.76 | 72.35±2.91 | 0.34 | 0.849 |
| 0.5<sup>b</sup> | 72.10±1.64 | 71.70±2.40 | 73.27±2.68 | 69.67±2.79 | 72.27±2.28 | 1.81 | 0.159 |
| 1<sup>b</sup> | 72.95±1.86 | 71.85±2.85 | 71.43±2.75 | 71.92±2.71 | 72.23±2.09 | 0.31 | 0.868 |
| 1.5<sup>b</sup> | 72.57±2.46 | 71.07±2.80 | 71.85±2.72 | 72.57±2.63 | 71.51±2.60 | 0.37 | 0.826 |
| 2<sup>b</sup> | 72.40±2.26 | 71.72±3.13 | 71.87±2.31 | 71.22±2.40 | 72.12±2.44 | 0.19 | 0.943 |
| 3<sup>b</sup> | 73.35±2.37 | 71.53±2.25 | 71.95±2.78 | 72.50±2.54 | 72.75±2.67 | 0.46 | 0.761 |

a: Before FDP injection; b:after FDP injection. P>0.05, No statistically significant difference between different dose groups at the same time point;
P < 0.05, with statistically significant differences between the different dose groups at the same time point.\*:P<0.01.

**Figure 6.**
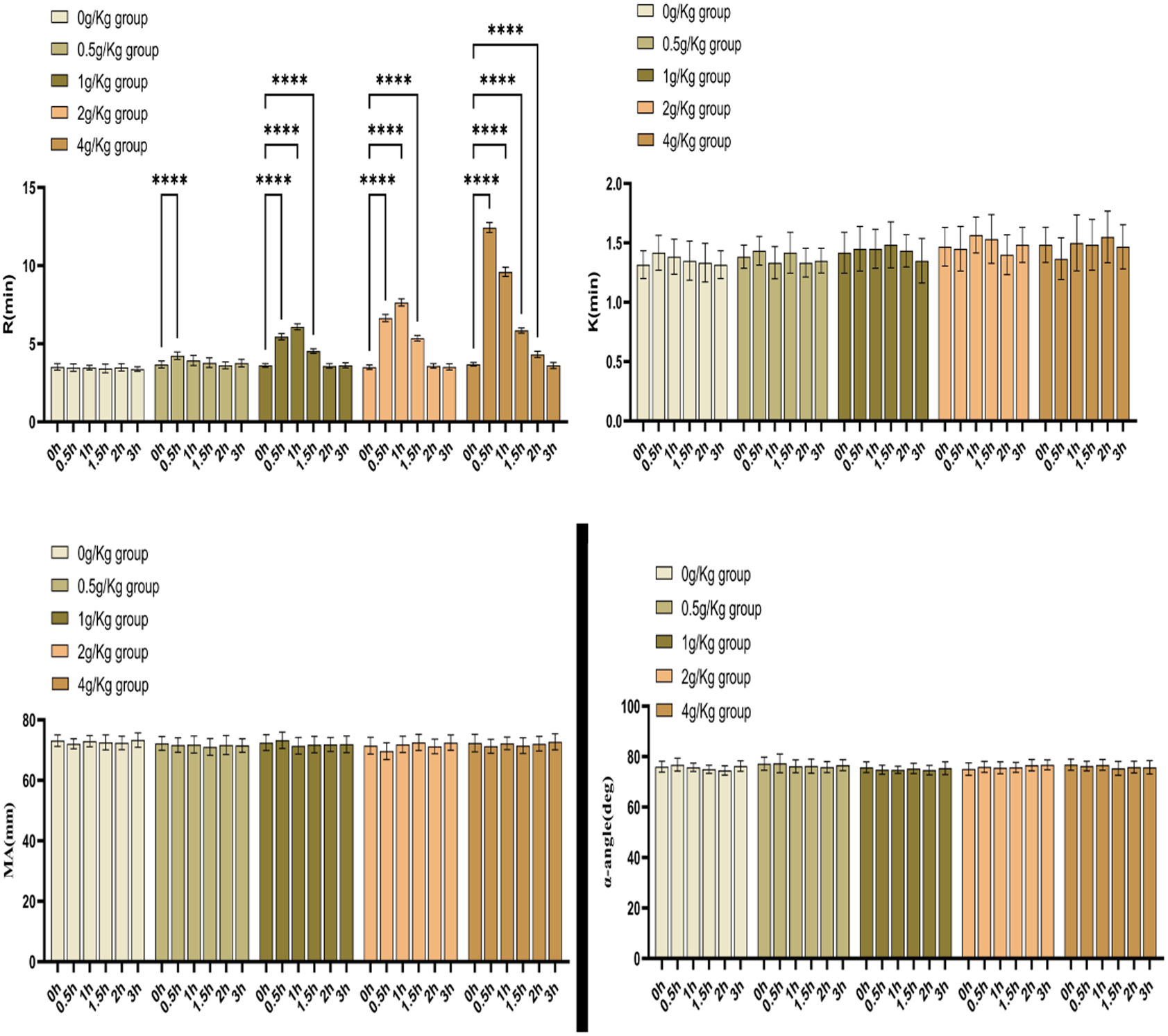
There were statistically significant differences between the R-values of the 0.5g/kg group at 0.5h post-dose, the 1g/kg group at 0.5h, 1h and 1.5h post-dose, and the 2g/kg group at 0.5h, 1h and 1.5h post-dose, and the 2g/kg group at 0.5h, 1h, 1.5h, and 2h post-dose and their R-values prior to the administration of the drug (P<0.0001); and there were no statistical differences (P>0.05) for the K, α -angle and MA values between time points compared with their pre-dose.

## 4. Discussion

To date, comprehensive literature reviews have revealed no prior reports regarding the effects of fructose-1,6-diphosphate (FDP) on coagulation reaction time(R) or coagulation factor activity^8,9^. Our research team serendipitously observed FDP-mediated alterations in thromboelastography (TEG) parameters (Maiketian Medical Systems) during routine laboratory investigations. To validate this phenomenon, we systematically replicated experiments using detection systems from Lepu Medical and Dingrun Biotechnology, confirming consistent results across three distinct platforms. This multi-platform verification enhances the reliability and reproducibility of our findings.

Mechanistic investigations demonstrated that FDP exerts concentration-dependent inhibitory effects on coagulation factors V, VII, IX, XI, and XII (p<0.001 for all), while maintaining negligible impact on factors II, VIII, and X. These findings correlate with TEG parameter changes: The R-time (clot initiation time, defined as the interval from blood sampling to 2mm amplitude on TEG tracing) showed dose-dependent prolongation with FDP exposure (p<0.001). As R-time reflects thrombin generation kinetics prior to fibrin polymerization[8], its extension suggests impaired early coagulation activation, consistent with observed factor activity suppression.

The established reference range for R in healthy adults is 5-10 minutes, primarily determined by coagulation factor levels and anticoagulant therapy status^9,10^. Our experimental data demonstrated FDP-induced R prolongation (Figure5), suggesting either direct inhibition of coagulation factor activity or interference with their activation processes. These findings were corroborated by parallel reductions in specific coagulation factor activities, establishing a coherent mechanistic explanation for the observed anticoagulant effects.

Clinical relevance was further explored through rabbit models. Intravenous FDP administration in New Zealand White rabbits (Oryctolagus cuniculus) produced dose-dependent R-time prolongation (Figure 5), mirroring in vitro findings. Although FDP is an endogenous metabolic intermediate, our data suggest therapeutic concentrations (1-6 mg/mL) may disrupt hemostatic balance. Notably, calculated human plasma concentrations following standard 5-10g IV dosing (1.86-3.71 mg/mL in 70 kg adults) fall within our experimental range, implying potential clinical implications.

To evaluate clinical relevance, we conducted pharmacodynamic studies in New Zealand White rabbits. Both in vitro and in vivo experiments demonstrated dose-dependent prolongation of coagulation parameters (p<0.001), indicating potential interference with early-stage coagulation initiation. While these findings suggest possible bleeding risk implications, it should be noted that FDP is an endogenous metabolic intermediate. Our experimental design focused on acute pharmacological effects, and comprehensive pharmacokinetic studies remain warranted.

Our findings extend previous reports of FDP-mediated prolongation of standard coagulation assays (PT, aPTT, TT)^11^. The observed concentration-dependent suppression of factors V, VII, IX, XI, and XII provides a plausible mechanism for these effects. This mechanistic convergence between conventional coagulation tests and thromboelastographic parameters strengthens the biological plausibility of FDP’s anticoagulant properties.The study extends previous work demonstrating FDP-induced prolongation of PT, aPTT, and TT^11^. Current results provide mechanistic clarity: Factor V/VII/IX/XI/XII inhibition directly explains these global coagulation test abnormalities. This concordance between specific factor assays and global coagulation parameters strengthens the biological plausibility of our findings.Pre-FDP administration coagulation testing Therapeutic drug monitoring in high-bleeding-risk populations Vigilance for perioperative bleeding tendencies

From a clinical perspective, FDP’s therapeutic applications in ischemic tissue protection^2,4,5^ necessitate careful consideration of its anticoagulant effects. Our in vitro data suggest that: (1) coagulation assays should ideally precede FDP administration, and (2) therapeutic monitoring may be prudent in patients with pre-existing coagulopathies. However, clinical translation requires caution given study limitations: (1) experimental concentrations (1-6 mg/mL) were extrapolated from theoretical human plasma levels (1.86-3.71 mg/mL for 5-10g IV dose in 70kg adults), (2) the exclusive use of in vitro models, and (3) lack of human pharmacodynamic data.

Clinical Implications as FDP demonstrates multifactorial benefits in ischemic tissue repair^2,4,5^ its coagulation-modulating properties necessitate cautious clinical application. We recommend: Pre-FDP administration coagulation testing: Therapeutic drug monitoring in high-bleeding-risk populations:Vigilance for perioperative bleeding tendencies:Future Directions Priority research areas include:Pharmacokinetic-pharmacodynamic modeling of FDP’s hemostatic effects, Prospective clinical trials assessing bleeding endpoints, Mechanistic studies exploring FDP-factor interaction modalities.

There are some limitations in this study. While in vitro concentrations mirror pharmacokinetic predictions, actual in vivo effects require validation through phase pharmacodynamic studies. The clinical bleeding risk associated with FDP-mediated coagulation changes remains unquantified.

In conclusion, our in vitro study confirmed that FDP has a clear and definite effect on our coagulation assay results, especially coagulation factor activity (V, VII, IX, Ⅺ, and Ⅻ) and coagulation reaction time . Whether FDP really affects our body’s coagulation function should be of great concern to us.

## Data Availability

All relevant data are within the manuscript and its Supporting Information files.

## Human and Animal Rights

All procedures performed in study involving human participants were in accordance with the ethical standards of the institutional and/or national research committee and with the 1964 Helsinki declaration and its later amendments or comparable ethical standards. The study was approved by our Institutional Review Board, Anhui No.2 Provincial People’s Hospital, China [(R) 2024-037].

## Ethical approval

The purpose of this study was to investigate the effects of FDP on coagulation reaction time and coagulation factor activity tests in vitro. This study was conducted in vitro and has no adverse effects on patients’ health, because the samples was randomly collected from 11 volunteers and experimenters did not have direct contact with volunteers. Demographic and clinical data were collected by a questionnaire. The the Ethical Committees of The Anhui No.2 Provincial People’s Hospital approved the study [protocol number :No. [(R) 2024-037] and all the participants provided their written informed consent in accordance with the Declaration of Helsinki.

## Acknowledgement

This work was supported by a fund from the Anhui Provincial Health Commission (Health Research Program of Anhui, AHWJ2023BAc20016; and Health Research Program of Anhui, AHWJ2023BAa20021).

## Authorship Contributions

**Conceptualization**: Tongqing Chen,Yalong Zhang.

**Conduct experiments**: Tongqing Chen, Xingguo Zhong.

**Formal analysis:** Yuan Fang.

**Funding acquisition:** Yuan Fang, Tongqing Chen.

**Resources:** Yuan Fang, Ning He.

**Perform data analysis**: Lin Zhou.

**Writing – original draft:** Tongqing Chen and Yuan Fang..

**Writing – review & editing:** Tongqing Chen.

## Data availability statement

All relevant data are within the manuscript.

## References

1. Zhang CS. et al. Fructose-1,6-bisphosphate and aldolase mediate glucose sensing by AMPK. Nature. 548(7665):112-116(2017).

2. Wang W, Liu M, You C, Li Z & Zhang YHP. ATP-free biosynthesis of a high energy phosphate metabolite fructose 1,6-diphosphate by in vitro metabolic engineering. Metab Eng. 42:168–174(2017).

3. Alva N, Alva R & Carbonell T. Fructose 1,6-bisphosphate : A summary of its cytoprotective mechanism. Curr Med Chem. 23(39) :4396–4417(2016).

4. Li TT, Xie JZ, Wang L, Gao YY & Jiang XH. Rational application of fructose-1, 6-diphosphate: From the perspective of pharmacokinetics. Acta Pharm. 65(2):147–157(2015).

5. Donohoe PH, Fahlman CS, B ickler PE, etal. Neuroprotection and in-tracellular Ca^2+^ modulation w ith fructose-1, 6-bisphosphate during in vitro hypoxia-ischem ia involves phospholipaseC-dependent signaling[ J]. B rain Res, 917(2): 158–166(2001).

6. Lancé M D. A general review of major global coagulation assays: thrombelastography, thrombin generation test and clot waveform analysis[J]. Thrombosis journal, 13(1): 1(2015).

7. Elliott, Andrea, Wetzel, Jeremy, et al. Thromboelastography in patients with acute ischemic stroke[J]. International journal of stroke: official journal of the International Stroke Society. 10(2):194–201(2015).

8. David WJ, James AD.TEG and ROTEM: technology and clinical applications[J]. Am J Hematol, 89(2):228–32(2014).

9. Snorre BS, Jerard S, Joar S, etal. The use of thromboelastography (TEG) in massively bleeding patients at Haukeland University Hospital 2008-15[J]. Transfus Apher Sci, 58(1):117–121(2019).

10. Kamal AH, Tefferi A, Pruthi RK. How to interpret and pursue an abnormal prothrombin time, activated partialthromboplastin time, and bleeding time in adults[J]. Mayo Clin Proc. 82(7):864–73(2007).

11. Chen TQ, Duan Chen D, Lu Chen L, et al. The effects of fructose diphosphate on routine coagulation tests in vitro[J]. Sci Rep, 12(1):304(2022).

